# Impact of a high-dose intensive-targeted motor rehabilitation programme integrating advanced technology for adults with central neurological conditions (INTeRAcT): protocol for a single-blind randomised controlled trial with a clinical, health economic and process evaluation

**DOI:** 10.1101/2025.10.13.25337718

**Authors:** M. Coremans, I. Allewijn, F. Bataillie, L. De Bruyn, M. Fobelets, F. Jacobs, S. Janssens, L. Pattyn, K. Putman, A. Schiltz, L. Tedesco Triccas, F. Van Thienen, G. Verheyden

## Abstract

**Introduction:** Chronic stroke and spinal cord injury are both associated with persistent motor impairments, resulting in reduced independence in daily life and lower quality of life. Although rehabilitation is essential to address these challenges, the amount of therapy provided during the chronic phase remains limited, while the long-term cost of care is substantial. Here, we describe the INTeRAcT protocol: a randomised controlled trial with a clinical, health economic and process evaluation, designed to assess the effectiveness of a high-dose, intensive-targeted and personalised rehabilitation programme.

**Methods and analysis:** In this single-blind randomised controlled trial, we aim to recruit 100 adults in the chronic phase after stroke or spinal cord injury. Participants will be randomised to either the INTeRAcT intervention group (n=50) or a control group receiving usual care (n=50). The intervention group will receive 90 hours of personalised motor rehabilitation over three weeks, including upper and lower limb therapy, with and without advanced technology, cardiovascular fitness training, and self-management education. After the intervention, both groups return to usual care and are followed up for nine months. Clinical outcomes will be assessed by a blinded assessor at baseline (T0), post-intervention (T1), and at nine-month follow-up (T2), with additional data collected throughout for the health economic and process evaluations.

The primary clinical outcome is independence in daily life, assessed using the Functional Independence Measure for participants with stroke, and the Spinal Cord Independence Measure for those with spinal cord injury. Secondary clinical outcomes include the EuroQol 5D-5L, Canadian Occupational Performance Measure, Goal Attainment Scaling, and the Fatigue Severity Scale. Additional stroke-specific secondary outcomes are the Action Research Arm Test, Fugl-Meyer Assessment, Functional Ambulation Categories, the 6 Minute Walk Test, 10 Meter Walk Test, and the Stroke Self-Efficacy Questionnaire. A multivariate linear model will be used to compare clinical changes between the two groups. Health economic data will be collected using diaries and questionnaires, capturing direct and indirect costs such as healthcare use, medication, transport, productivity loss, and informal care. Effects will be measured via the EQ-5D-5L and expressed in Quality-Adjusted Life Years. Cost-effectiveness will be assessed through a trial-based cost-utility analysis over a nine-month horizon and a Markov model over a lifetime horizon, complemented by scenario analyses and a budget-impact analysis.

The process evaluation, guided by the UK Medical Research Council framework, adopts a mixed-methods approach combining quantitative and qualitative data from structured diaries, semi-structured interviews, and observations. Quantitative data will be presented descriptively, while qualitative data will be examined using both content and thematic analysis.

**Ethics and dissemination:** Ethical approval was obtained from the central and local ethics committees on 10 July 2023. The currently approved protocol is version 5, dated 15 March 2024. All participants provide written informed consent prior to enrolment. Study findings will be disseminated through peer-reviewed journals and presentations at national and international conferences, as well as at various stakeholder events.

**Trial registration number:** NCT05970367 (31 July 2023)

**ARTICLE SUMMARY:** *Strengths and limitations of this study:*

- This protocol combines clinical, process, and health economic evaluations to comprehensively assess a complex, high-dose and individually tailored motor rehabilitation intervention, addressing key evidence gaps regarding effectiveness, implementation, contextual factors and cost-effectiveness in chronic stroke and spinal cord injury care.
- Clinical outcome assessments will be conducted by a blinded assessor, reducing the risk of measurement bias.
- The rehabilitation programme combines a standardised structure with individual goal-setting, developed in collaboration with clinical practice and grounded in evidence-based principles, enhancing feasibility and clinical applicability.
- Although the trial is conducted in a single clinical centre, the inclusion of both stroke and spinal cord injury populations enhances generalisability across neurological conditions, while the process evaluation will identify contextual factors to support future implementation in other settings.

## INTRODUCTION

Globally, one in four people over the age of 25 will experience a stroke in their lifetime, resulting in nearly 12 million new strokes each year[1]. In 2021, there were 1.74 million incident stroke cases and 13.4 million prevalent stroke cases reported in Europe[1]. Predictions indicate that by 2030, an additional 3.5% of the adult population will have experienced a stroke[2]. For spinal cord injury (SCI), the incidence in Europe was estimated at 108,000 new cases in 2019, with 2.67 million individuals living with SCI[3].

Both conditions contribute to a significant disease burden[1,4] affect multiple domains of human functioning[5] and are associated with a reduced quality of life[6,7]. Motor problems are the most common impairment[8,9], affecting 60–75% of patients after stroke[9] and even more than 80% of individuals with SCI[10], even in the chronic phase[10,11]. Thus, despite completing standard rehabilitation in the subacute phase, deficits remain present in a large number of patients. Motor impairments can involve both the upper and lower limbs, leading to significant limitations in locomotion[12,13] and dependency during daily functioning[14]. To date, there is no cure for post-stroke and post-SCI deficits, but rehabilitation can effectively address motor impairments such as reduced strength and balance, as well as functional limitations in performing activities of daily living.

In the literature, the so-called “critical window” describes a period of heightened neuroplasticity during which the greatest motor recovery occurs, typically within the first three months following a stroke or SCI[15–19]. Around six months post-onset, motor recovery tends to plateau and generally remains below pre-pathology function[15–19]. However, Ballester et al.[20] demonstrated that an increased sensitivity to rehabilitation can persist beyond one year after stroke. A similar phenomenon has been observed in chronic SCI, where several studies have reported motor improvements more than one year after onset[21–23]. These findings suggest that patients retain the capacity to enhance their functional abilities through exercise-dependent plasticity[24,25]. Despite this ongoing potential for improvement in the chronic phase, rehabilitation services during this phase remain limited within the current healthcare system.

Increasing the dose of rehabilitation provides beneficial effects, such as greater improvement in motor ability, reduced activity limitations, and enhanced functional performance[26–28]. Similarly, the integration of advanced rehabilitation technology with conventional therapy has proven effective in improving motor function[29–32] and promoting motor learning[33,34]. In this context, Ward et al.[25] provided an intensive rehabilitation programme for the upper limb, combining patient-centred therapy with repetitive movements using robotic devices, for individuals with chronic stroke. The programme totalled 90 hours, delivered over three weeks with six hours of therapy per day, five days a week. A total of 224 patients completed the programme, resulting in significant and clinically meaningful improvements in upper-limb impairment and activity, which were maintained at six-month follow-up. Although intensive rehabilitation programmes often yield positive results, they still present methodological limitations, such as the absence of a control group and the focus on a single body region or therapy component (e.g., upper limb rehabilitation or isolated use of technology)[25,27,35]. Building on the study of Ward et al.[25] who demonstrated the effectiveness of high-dose upper limb rehabilitation, we will implement a high-dose, intensive-targeted motor rehabilitation programme for both the upper and lower limbs. This 90-hour programme integrates conventional physiotherapy with advanced technology, cardiovascular fitness training, patient-centred rehabilitation, and self-management education.

To make this level of intensity feasible and effective, advanced rehabilitation technologies will be employed. Beyond their demonstrated effectiveness in improving motor function[29–32] and quality of life[36], these technologies offer several advantages. They allow for a greater number of repetitions within a given time frame without overburdening therapists[37], enable the safe progression of intensity[38], and promote patient engagement through gamification, which enhances motivation and supports adherence to recommended therapy doses[39]. These advantages are particularly relevant for chronic stroke and SCI patients, for whom the perception of limited therapy tolerance has been disproven by studies showing successful completion of 300 hours of upper limb therapy over 12 weeks[35,40].

Cardiovascular fitness training is both safe and effective for improving aerobic capacity[41,42]. It plays a crucial role in promoting long-term health and enhancing the quality of life in individuals with SCI[43–46] and stroke[42,47–50]. In addition to improving physical fitness, regular cardiovascular exercise helps prevent cardiovascular disease[51,52] and serves as a strategy for secondary stroke prevention[53].

Person-centred goal setting is a fundamental aspect of neurological rehabilitation, yet it remains underused[54,55]. When applied effectively, it enhances a patient’s self-confidence, motivation, treatment adherence, and engagement in rehabilitation[55–57]. Evidence suggests that involving patients in goal setting increases satisfaction with their rehabilitation experience and the achievement of personally meaningful outcomes[58]. Additionally, goal achievement has been associated with improved self-efficacy and more positive perceptions of participation in daily and community life[59] These benefits contribute to a more rewarding rehabilitation process, promoting long-term success.

Lastly, a self-management programme is included to equip participants with the knowledge and skills to manage their symptoms and the impact of their condition on daily functioning[60]. Self-management has been shown to reduce the negative consequences of chronic conditions and to improve overall health and quality of life[61]. Strengthening self-management skills may also positively influence rehabilitation outcomes, functional independence, and participation[62,63].

Therefore, the primary aim of this study is to conduct a phase III clinical trial, including a high-quality randomised controlled trial (RCT), to evaluate the clinical effectiveness of this novel motor rehabilitation package.

One of the main cost drivers in long-term health care is the level of disability[64,65]. In addition, the increasing incidence and prevalence of stroke and SCI, driven by different underlying reasons such as increased exposure to risk factors, improved survival rates and shifting demographic trends, will substantially increase the economic burden of these conditions across Europe[3,66–70]. This translates into increased expenditures for both healthcare and social services[71,72]. For stroke, the total cost of ongoing care is estimated at €45 to €60 billion annually[66] and is expected to rise further over the next decade[73]. Although less prevalent, SCI also imposes a considerable financial burden, with estimated annual costs ranging from €92 to €212 million[72]. Neurorehabilitation is resource-intensive, therefore, it is essential to consider the potential trade-offs between relevant costs and effects of rehabilitation care. However, a persistent knowledge gap between clinicians, health economists, and policymakers hampers the development and implementation of innovative rehabilitation approaches. This is partly due to the limited, inconsistent, or inconclusive nature of existing studies[74] in the emerging field of “rehabilitation economics”. While several studies have suggested that increased rehabilitation intensity in stroke may be beneficial, the overall evidence remains inconclusive[75]. For spinal cord injury, economic evaluations have primarily focused on acute rehabilitation or locomotor-specific interventions[76–78]. Moreover, few studies have investigated the cost-effectiveness of advanced technology-based rehabilitation[74]. Although some interventions demonstrated cost-effectiveness or reductions in healthcare utilisation[79–82], most studies were limited by too short follow-up periods[81] or by the use of a single technological device[79,81,83].

To accurately evaluate the cost-effectiveness of new rehabilitation approaches in the chronic phase, there is a need for clinical trials conducted in outpatient settings[84,85], as well as economic evaluations performed from both the healthcare system and societal perspectives[86]. Additionally, employing micro-level data, where all resources utilised by each individual participant are directly recorded, can significantly enhance the accuracy and reliability of cost estimations[87,88].

Therefore, a second aim of this study is to conduct a health economic evaluation alongside the INTeRAcT clinical trial, both to contribute to the existing evidence and to support policymakers in making informed decisions regarding the clinical implementation and potential reimbursement of our INTeRAcT programme.

Given the complexity of rehabilitation interventions, multicriteria decision analysis may facilitate decision-making by incorporating a broader spectrum of evaluation criteria beyond traditional cost-effectiveness and clinical efficacy metrics. These may include stakeholder experiences (e.g., patients and therapists), contextual factors, feasibility, reproducibility, and long-term sustainability[89]. In parallel, the implementation of novel, evidence-based interventions can be associated with numerous challenges[90]. Consequently, a process evaluation is considered essential when evaluating complex interventions[91,92], as it provides critical insights into implementation quality and fidelity, helps to uncover causal mechanisms, and identifies contextual influences that may explain variations in outcomes[93,94]. Additionally, such evaluations are valuable in understanding why interventions may fail or yield unintended consequences, and in identifying ways to optimise them[93,94].

For instance, in the Queen Square programme by Ward et al.[25], the process evaluation highlighted that the psychosocial aspects of the intervention were perceived as equally important as the intensity and behavioural training by stroke survivors, their caregivers, and clinicians[95].

Hence, the third objective of this study is to conduct a process evaluation of the INTeRAcT programme, to inform and support its implementation into clinical practice and health policy.

In summary, the overall aim of this randomised, single-blind controlled trial is to evaluate the clinical effectiveness and cost-effectiveness of the INTeRAcT programme, a high-dose intensive-targeted 90-hour motor rehabilitation programme integrating advanced technology, in adults with stroke or spinal cord injury, alongside conducting a process evaluation.

We hypothesize that the INTeRAcT programme will (I) lead to significantly greater improvements in functional independence at both immediately post-intervention and nine months post-baseline; (II) result in clinically relevant improvements in patient-relevant goals, fatigue and quality of life; (III) yield significantly greater improvements on the stroke-specific outcome measures; and (IV) be cost-effective compared, to usual care. In addition, the process evaluation will provide valuable insights to inform and guide future implementation of the INTeRAcT programme in daily clinical practice.

## METHODS

### Study design

The INTeRAcT trial is an assessor-blinded, randomised, phase III controlled trial (Figure 1). A total of 100 participants with stroke or spinal cord injury (SCI) will be randomly assigned to either a three-week rehabilitation programme (INTeRAcT, n=50) or a control group receiving usual care (n=50). Participants will be stratified for condition and earlier rehabilitation experience with technology. Following the initial three weeks, both groups will continue their usual care. All participants will be assessed by a blinded assessor at three time points, before the intervention (T0, baseline), after three weeks (T1), and nine months post-baseline (T2); and will be followed up for nine months after baseline. Parallel to the clinical study, a process evaluation and a health economic evaluation are conducted.

**Figure 1.**
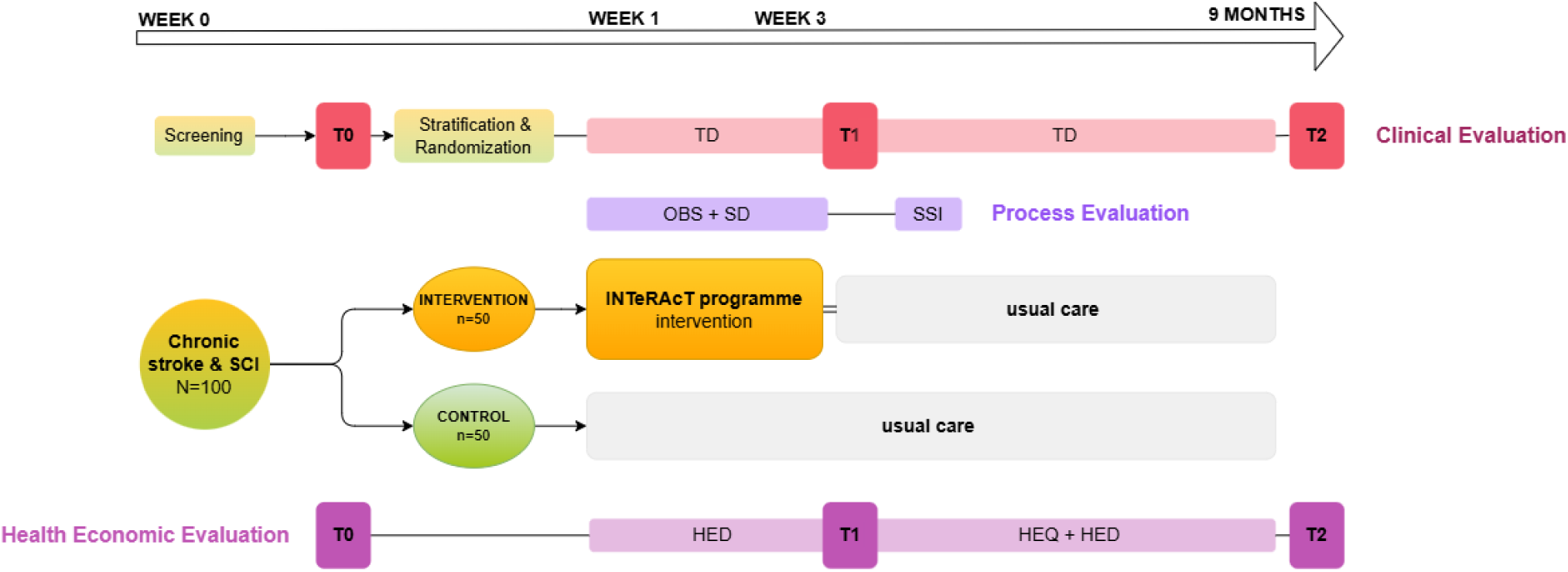
The INTeRAcT study design: The study includes outcome assessments at three time points (T0, T1, and T2), a 3-week intensive rehabilitation programme for the intervention group, and a 9-month follow-up period. In parallel with the clinical evaluation, both a process evaluation and a health economic evaluation are conducted. TD = therapy diary; OBS = observation; SD = structured diaries; SSI = semi-structured interviews; HED = health economic diaries; HEQ = health economic questionnaires.

### Participant selection and recruitment

In this RCT, adults in the chronic phase after a stroke or SCI, living in Belgium, participated. Recruitment was conducted via databases (Stroke Rehabilitation Research Team, KU Leuven/To Walk Again), information sessions in hospitals and rehabilitation centres, and outreach to participant associations and adaptive sports centres in Belgium. First-line healthcare providers, including general practitioners, physiotherapists, speech therapists, home care nurses, and pharmacies, were encouraged to inform potential participants.

The inclusion criteria were: (1) First stroke or spinal cord injury (ASIA A, B, C, or D) with a clear participant need related to the upper and/or lower extremities, diagnosed by a medical specialist more than 3 months ago; (2) living at home and more than 3 months post-discharge from a hospital or rehabilitation centre; (3) a maximum recovery of 85% independence in daily functioning, assessed using the Functional Independence Measure (FIM) for stroke or the Spinal Cord Independence Measure (SCIM) for SCI; (4) a pre-pathology Barthel Index >85/100; (5) unilateral or bilateral weakness of the affected upper and/or lower limbs, with no maximum score of 5 on all three segments of the Motricity Index; (6) age ≥18 years; and (7) clinical and cognitive ability to perform high-dose rehabilitation with advanced technology, as assessed by a medical doctor (MD).

Exclusion criteria were: (1) no voluntary movements against gravity in the upper and lower limbs; (2) other neurological or musculoskeletal conditions that could interfere with protocol adherence, as assessed by an MD; (3) severe communication, cognitive, language, or visual impairments preventing participation in the intervention or assessment process, as assessed by an MD; (4) any condition that could compromise participant safety or compliance with the clinical investigation plan, as determined by the investigator and MD; and (5) pregnancy or breastfeeding. Trial information was provided, and informed consent was obtained by a researcher of KU Leuven before screening. The screening was conducted by a researcher from KU Leuven in collaboration with a rehabilitation physician from AZ Herentals.

### Sample size calculation

The power calculation was performed based on the primary research question: the evaluation of the short-term clinical effect of the INTeRAcT intervention versus usual care at home on independence in daily living, measured using the FIM/SCIM (see below: clinical evaluation). Since no previous studies have assessed functional independence in both stroke and spinal cord injury participants in the chronic phase as a combined group, the calculation was based on a relevant improvement for this mixed population. A total sample size of 90 participants will provide 80% power to detect a 12% significant and relevant difference[96,97] in FIM/SCIM scores between the two groups, assuming a standard deviation of 20%[98,99] and a two-tailed alpha of 0.05. This corresponds to an effect size of 0.6, which is considered a “medium” effect. To account for an expected dropout of 10%, 100 participants will be recruited.

### Randomisation and blinding

To ensure the integrity of the study, the following randomisation procedures have been implemented. Participants (n=100) are randomly assigned to one of two groups: the intervention group, “INTeRAcT” (n=50), or the control group, “usual care” (n=50). To achieve a balanced distribution across conditions and prior rehabilitation technology experience, participants are stratified by condition (stroke or SCI) and prior technology experience. The latter refers to previous participation in outpatient rehabilitation involving advanced technological devices at a frequency of at least one to two sessions per week for a minimum duration of three months. Allocation concealment is ensured, with randomisation lists prepared in advance using a block size of two. Randomisation will be performed by a member of the study team who is not involved in participant contact or data collection. Due to logistical restrictions, a predefined quota has been set for groups of five participants, with three allocated to the intervention group and two to the control group. Participants will be randomly assigned to one of the two groups as they enter the study. Midway through recruitment, the allocation ratio will shift to 3 participants in the control group and 2 in the intervention group, ensuring balanced group sizes.

To ensure objectivity and minimise bias, outcome assessments will be conducted by an assessor who is blinded to treatment allocation. The group assignment will be revealed to participants after the first measurement (T0). In exceptional cases, due to practical arrangements for participation in the 3-week intervention, the group may be revealed to the participant earlier, however, the assessor will remain blinded. Participants will be instructed not to disclose their group allocation. The physiotherapists, participants after T0 and researchers conducting follow-up, process and economic evaluation cannot be blinded to allocation.

### Trial treatment

#### Experimental intervention: INTeRAcT programme

The intervention group will receive a total of 90 hours of motor therapy over three weeks, 30 hours per week, delivered in six-hour sessions each weekday. The therapy will be provided by trained physiotherapists with expertise in neurological disorders at the clinical site, AZ Herentals (To Walk Again). This high-dose, intensive-targeted rehabilitation programme is evidence-based, grounded in the principles of motor learning, personalised, and goal-oriented. The programme aims to enhance physical fitness and independence in daily life through cardiovascular fitness training, therapy for the upper and lower limbs, with and without advanced technological devices, and goal-oriented training. All of this is tailored to the individual participant, considering personal goals and limitations, incorporating built-in progression over time. Additionally, the programme emphasises transferring skills to functional tasks and promoting self-management to improve the participant’s self-efficacy beyond the duration of the programme. These components constitute the seven therapy blocks of the programme. All blocks are covered daily in a random order, except cardiovascular fitness training and self-management. Figure 2 illustrates an example of a weekly schedule that outlines the various therapy components.

**Figure 2.**
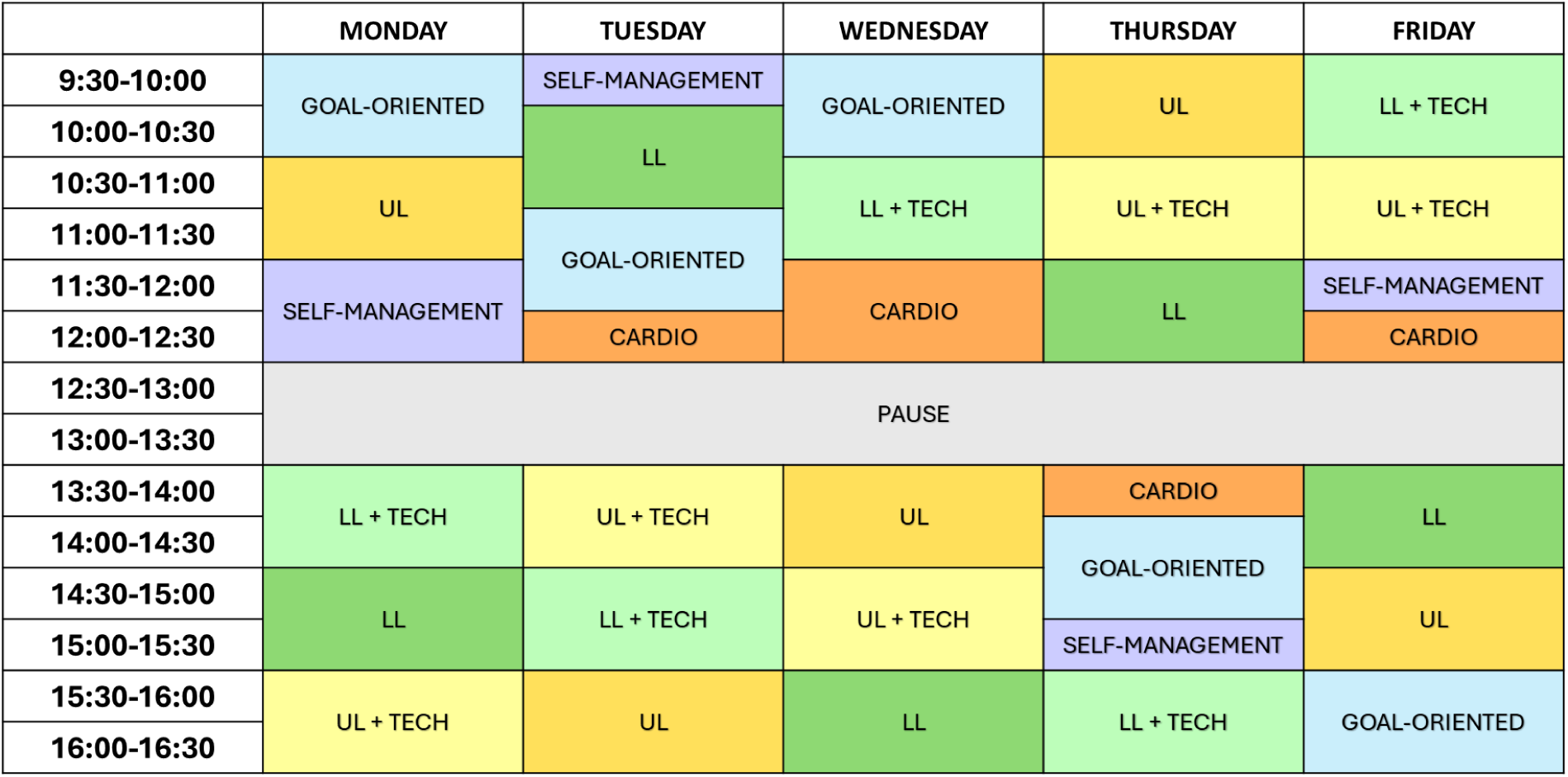
Example of the intensive programme for one week: The therapy programme is structured in daily six-hour sessions, comprising the following seven components: (1) upper limb therapy (UL), (2) lower limb therapy (LL), (3+4) therapy with technological devices for the upper or lower limb (TECH), (5) cardiovascular fitness training (CARDIO), (6) goal-oriented therapy, (7) self-management education.

During the baseline (T0) assessment, three specific goals will be defined by the participant, in collaboration with a KU Leuven researcher with a master’s degree in neurological physiotherapy, using the Goal Attainment Scaling (GAS)[100,101], which will serve as the focus throughout the three-week intervention. Based on these goals, the physiotherapist will evaluate which subgoals are still needed to achieve the goal. The overall structure of the programme follows a four-step process:

1. Setting personalised goals.
2. Evaluating and observing the activity to identify the subgoals contributing to the main problem.
3. Practising these subgoals to address underlying difficulties during components 1-5, as outlined below.
4. Practising the full activity during the goal-oriented therapy sessions (component 6), as outlined below.

Upper and lower limb therapy sessions without technology (components 1 & 2) follow a conventional therapy approach and include a range of techniques such as mobilisation and stretching of the affected limb, analytical and coordination exercises, strength training, task-specific training with repetitions, fall prevention, balance training, and functional exercises. These sessions incorporate both proximal and distal components, emphasising fine manipulation, sensorimotor training, foot placement, analytical movements, gait adaptability, and complex activities of daily living. Both upper and lower limb therapy components are provided daily, one hour each, for a total of five hours per week per component.

In addition to conventional upper and lower limb therapy, participants participate in technology-assisted upper and lower limb therapy (components 3 & 4), with one hour per day dedicated to each component. For upper limb rehabilitation, the Gloreha (BTL Robotics, Belgium) focuses on improving distal hand function through passive or active-assisted mobilisation of the fingers. Action observation and interactive games are integrated to stimulate arm and hand use while providing arm support. The OmniHi5™ (ACP, USA) uses electrical stimulation to facilitate upper limb movement. The HUR fitness equipment (HUR, Finland) is adapted for individuals in wheelchairs for strength training, but can also be used by ambulatory individuals with a standard chair. For lower limb rehabilitation, an Exoskeleton gait robot (Ekso Bionics, USA) assists individuals with limited or no walking ability, whereas the ZeroG Gait & Balance System (Aretech, USA) provides dynamic body weight support in a fall-safe environment to facilitate gait training. The RehaMove (Hasomed, Germany), a Functional Electrical Stimulation bike is used to activate muscles and promote movement. Additionally, the DIERS 4D motion® Lab (DIERS, Germany) integrates a treadmill with a feedback system to improve weight shifting and promote a more normal gait pattern. To complement these technologies, the D-WALL Elite (TecnoBody, Italy) offers a versatile platform for designing personalised exercises and managing a broad range of programs focused on posture, functional training, balance, and strength. These advanced technologies are integrated into therapy to align with the participant’s individual rehabilitation goals.

The cardiovascular fitness training[102–104] (component 5) focuses on interval training, using a variety of methods such as a bicycle/arm ergometer, stepping, and TABATA (e.g., boxing, circuit training, ropes). The training intensity is primarily monitored using heart rate. In individuals with complete or incomplete spinal cord injury, where muscle mass is limited, the intensity is guided by the Borg Rating of Perceived Exertion Scale (BORG PRE)[105], aiming to achieve a score of at least 12 out of 20. This training is delivered in a group setting for a total of 2.5 hours per week, consisting of one 1-hour session and three 0.5-hour sessions.

During the goal-oriented training session[106] (component 6), the therapist and participant collaboratively design and execute an individualised therapy plan based on the three specific goals previously established. These goals follow the SMART principles (Specific, Measurable, Achievable, Relevant, Time-bound). For example, a goal might be: “After three weeks of training, I want to be able to walk 2 km with my dog”. This approach ensures that therapy is tailored to the participant’s needs and integrated into a highly functional, real-life context. The goal-oriented training is conducted daily, with one hour of individual sessions each day, amounting to a total of five hours per week.

The self-management sessions (component 7) are based on key principles that emphasise a personalised and flexible approach. It tailors support to individual needs and circumstances, allowing for initiation at any stage. The approach prioritises the person’s story and promotes supportive relationships while encouraging autonomy. Emphasis is placed on small, meaningful actions in daily life, reinforcing autonomy and motivation. It builds on individuals’ existing self-management strategies and social support systems to enhance long-term engagement and adaptability[107]. Self-management sessions comprise a total of 2.5 hours per week, including one hour of group session and three individual sessions of 0.5 hours each. Group therapy allows participants to learn from each other and exchange experiences and strategies, while individual sessions provide personalised counselling tailored to each participant’s needs and goals.

During the three-week intervention period, the intensive therapy is documented daily in diaries by the physiotherapists, including the total number of minutes of therapy, the division between active therapy time (e.g., performing active repetitive movements) and other therapy time (e.g., explanation, resting periods, activities of daily Living (ADL) tasks), and the content of each of the seven therapy blocks. Additionally, at the end of each therapy block, the BORG PRE[108] is completed to assess participants’ subjective experiences of physical exertion. This scale ranges from 6 (“no exertion at all”) to 20 (“maximal exertion”) and allows for individualised adjustments to training intensity. Outside the 6-hour daily intervention sessions, participants were permitted to continue their usual care, which was recorded in diaries.

#### Control intervention: usual care

To allow a pragmatic comparison with current clinical practice, the control group will continue their usual care at home or in an outpatient rehabilitation centre or hospital for the entire 9-month study period, without receiving any alternative interventions. After completing the study, participants in the control group will be given the opportunity to receive the identical 3-week programme following the final follow-up measurement at 9 months. This waitlist-control design will help maintain participants’ motivation to adhere to the study during the 9-month follow-up period.

Participants will document their usual care in therapy diaries (Figure 1), with assistance from their therapist if needed. These diaries will capture the frequency (number of sessions), intensity, time (duration), and type (therapy content) of physiotherapy, occupational therapy, and psychological sessions, in line with the FITT principles[104]. The diaries were developed by the research team based on expert input, and their content was reviewed and approved by clinicians. Participants in the intervention group will complete the same diaries during the follow-up period, after the 3-week intervention. The diaries will be collected and reviewed monthly, and any missing data will be requested from participants.

### Adverse events

The risk of adverse events (AEs) occurring during this study is considered low, therefore, safety reporting will be limited to the safety reporting that is necessary in routine care. Participants will be asked to report any AE related to the study-specific intervention to the study team. The study team will keep detailed records of all reported AEs, which will be assessed by the investigator at the rehabilitation site, a medical doctor, regarding their seriousness, causality, and expectedness. A structured procedure has been established to ensure proper reporting of AEs to the relevant regulatory committees. If an AE directly or indirectly related to study participation requires additional medical care, the associated costs will be covered by the study’s insurance.

The following events are commonly observed and are therefore not considered as AEs for the study: (1) During or after the treatment, the participant may feel the following symptoms or events: fatigue, muscle spasm/increase in muscle tone and soreness, as with any movement exercise with a device that activates the body and muscles. These types of symptoms are anticipated and should fade out; (2) Redness of the skin, light bruising or small superficial pressure injuries where the equipment has exerted pressure on the skin, from harnesses or use of the revalidation robots. These injuries will be subject to follow-up as they are in usual care: close monitoring by the therapist and use of pressure bandages to prevent them; (3) Orthostatic hypotension: blood pressure drop due to position change (usually due to verticalizing: sitting/lying to standing); (4) Changes in bladder and bowel control after activation of the participant.

Premature discontinuation or interruption of the intervention may occur due to AEs, protocol violations, pregnancy, or illness. In such cases, clinical follow-up will be ensured, and if illness interferes with therapy, continuation or restarting the intervention will be considered where feasible.

### Participant and public involvement

Survivors of stroke and spinal cord injury reviewed and provided input on the various study documents that participants are required to complete during the study. Adjustments were made based on their feedback. A scientific committee composed of clinical stakeholders and researchers has been established and will be involved throughout the entire course of this study.

### Outcomes

#### Clinical evaluation

The primary objective of this study is to evaluate the clinical benefits of the INTeRAcT programme across various domains of the International Classification of Functioning, Disability, and Health (ICF) immediately post-intervention and after nine months of follow-up[109]. Therefore, participants are assessed by a blinded assessor at three different time points: (T0) baseline, (T1) immediately post-intervention, and (T2) nine months post-baseline (Figure 1). The assessor remains blinded to treatment allocation to ensure objective outcome evaluation. Participants will be reminded of their upcoming assessment appointments in a timely manner via email or telephone. During data collection, a standardised case report form (CRF) is completed. This form includes researchers’ notes, demographic and pathology-related data, and the results of the standardised clinical assessments and questionnaires.

##### Data: clinical outcome measures

Different outcome measures are collected by a trained, blinded assessor, as shown in Figure 3. At baseline (T0), demographic, socio-economic, and health-related data will be collected. An overview is provided in Appendices 1A and 1B.

**Figure 3.**
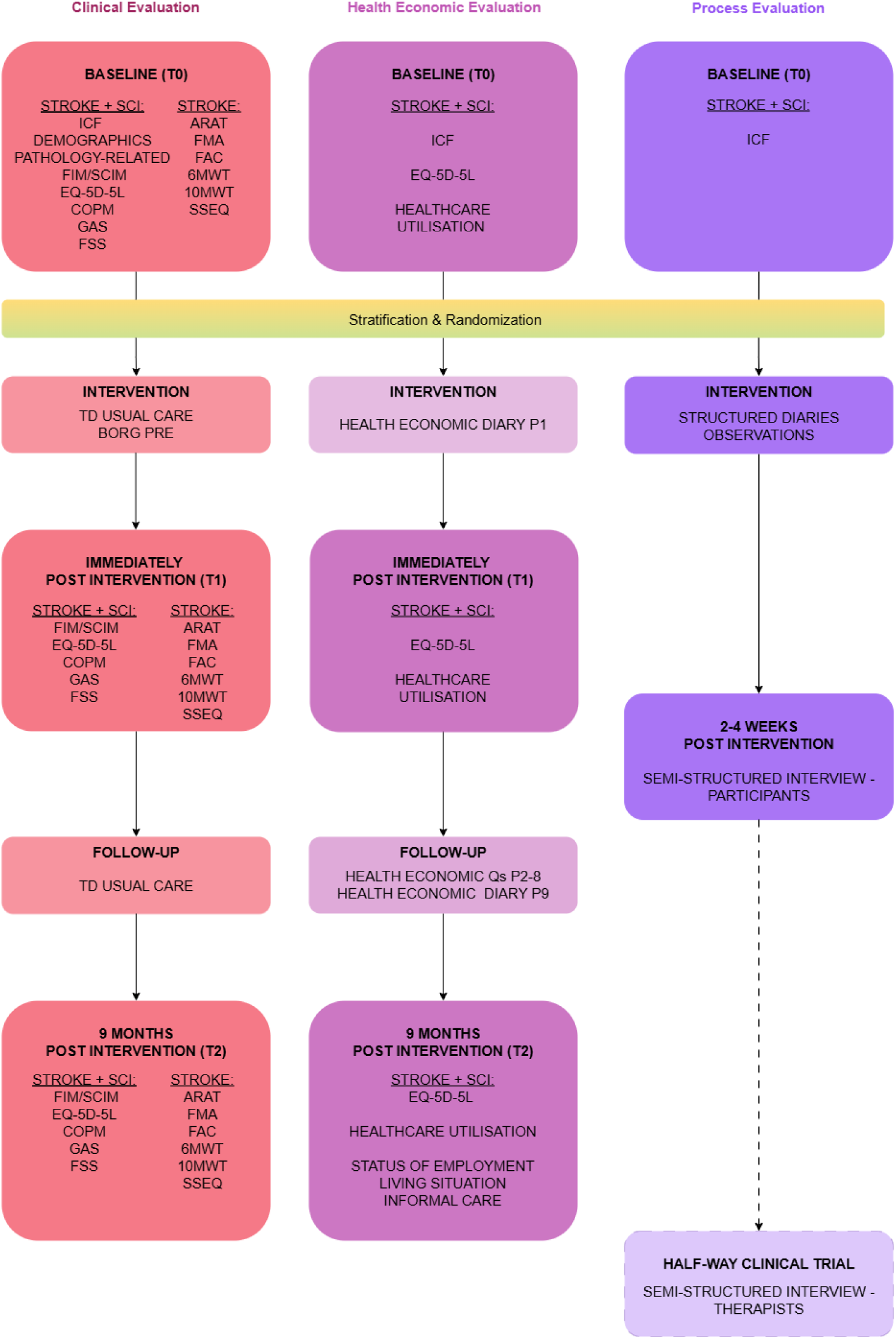
Flowchart for data collection of the INTeRAcT trial. The figure outlines the timing of clinical, health economic, and process evaluations for participants throughout the study. The “halfway clinical trial” section indicates data collection of the interviews with the INTeRAcT therapists. FIM = Functional Independence Measure; SCIM = Spinal Cord Independence Measure; EQ-5D-5L = EuroQol 5-Dimensions 5-Levels; COPM = Canadian Occupational Performance Measure; GAS = Goal Attainment Scaling; FSS = Fatigue Severity Scale; ARAT = Action Research Arm Test; FMA = Fugl-Meyer Assessment; FAC = Functional Ambulation Categories; 6MWT = 6-Minute Walk Test; 10MWT = 10-Metre Walk Test; SSEQ = Stroke Self-Efficacy Questionnaire; BORG PRE = Borg Rating of Perceived Exertion Scale; TD = therapy diary; Health economic Qs = health economic questionnaires

The primary outcome of this study is the change in functional independence from baseline to T1 (post-intervention), assessed using the Functional Independence Measure (FIM) and the Spinal Cord Independence Measure (SCIM)

> **The FIM-motor**[110] evaluates independence in daily activities following a stroke. It comprises 13 items that assess the level of assistance required for performing activities of daily living safely and efficiently. These activities include self-care, bladder and bowel management, transfers, and mobility, and are scored on a 7-point ordinal scale ranging from total assistance to complete independence, with a total score between 13 and 91. The FIM has demonstrated acceptable reliability and validity in individuals with stroke[111,112].
>
> **The SCIM**[113] assesses daily functional achievements in individuals with spinal cord lesions. It consists of 19 items divided into three subscales: self-care, respiration and sphincter management, and mobility. The total SCIM scores range from 0 to 100, with 0 indicating total assistance and 100 indicating complete independence. The SCIM has been validated and is a reliable measure for this population[114].

To compare functional improvements across both participant groups, the percentage of improvement in FIM and SCIM scores will be analysed. Since the primary outcome measure for both participants after stroke and spinal cord injury is a functional improvement, the same construct applies to both populations. Both measures will be administered at T0, T1, and T2.

We include several secondary outcome measures representing different levels of the ICF for both stroke and SCI. All secondary outcomes will be administered at T0, T1, and T2.

> **The EQ-5D-5L**[115] measures health-related quality of life across five dimensions: mobility, self-care, daily activities, pain/discomfort, and anxiety/depression. Each dimension has 5 response levels: no problems, slight problems, moderate problems, severe problems, and unable to/extreme problems. A weighted health state index is calculated for individuals ranging from less than 0 (a health state perceived as worse than death) to 1 (representing full health), with higher scores indicating better health utility. Additionally, the questionnaire includes a visual analogue scale, where participants rate their perceived health from 0 (worst imaginable health) to 100 (best imaginable health). This scale applies to both stroke and SCI populations[116,117].
>
> **The Canadian Occupational Performance Measure (COPM)**[118] is an evidence-based outcome measure designed to capture a participant’s self-perception of performance in everyday living over time, across three areas: self-care, productivity, and leisure. The participant provides a score between 0 to 10 for importance, performance, and satisfaction. An average score for performance and satisfaction is calculated across all relevant items. It can be used in both study populations[119,120]. In this study, the COPM will also be used to assess the participants’ needs and establish the goals for the Goal Attainment Scaling, as described below.
>
> **The Goal Attainment Scaling**[100] is an individualised evaluation method scored on an ordinal 5-point scale, capturing a person’s individual treatment goals and the extent to which they are achieved. Each goal is rated from +2 (much more than expected) to -2 (much less than expected), with 0 representing the expected level of achievement. The baseline level is typically set at -1, and goals can be weighted by the participant according to their importance or difficulty. GAS will be used for both stroke and SCI populations[121].
>
> **The Fatigue Severity Scale (FSS)**[122] is a 9-item scale, assessing the perceived severity of fatigue symptoms over the past week in various daily situations. It evaluates how fatigue impacts functioning and is scored on a 7-point scale, ranging from “strongly disagree” to “strongly agree,” with a minimum total score of 9 and a maximum of 63, with a higher score indicating more impact on daily life. It applies to both populations[123,124].

For the stroke population, additional secondary outcome measures were added, representing different levels of the ICF. All these secondary outcomes will be administered at T0, T1, and T2.

> **The Action Research Arm Test (ARAT)**[125,126] is an observational measure to assess upper extremity performance, including coordination, dexterity, and functioning. It consists of 19 items, with task performance rated on a 4-point scale, ranging from 0 “no movement” to 3 “movement performed normally”, resulting in a total score between 0 and 57, with higher scores indicating better upper limb performance.
>
> **The Fugl-Meyer Assessment (FMA)**[127,128] assesses motor functioning in the upper and lower limbs in post-stroke participants. It includes 50 test items, with a 3-point ordinal scale for each (0 = unable, 1 = partial, 2 = (near) normal), resulting in a score between 0 and 100, where 0 indicates no motor function and 66 indicates an intact motor function.
>
> **The Functional Ambulation Category (FAC)**[129,130], a 6-point functional walking test that evaluates ambulation ability, determining how much human support a participant requires when walking and whether or not they use a personal assistive device. A score of 0 indicates the participant cannot walk or needs help from two or more persons, while a score of 5 indicates the participant can walk independently.
>
> **The 6-minute Walk Test (6MWT)**[131,132] measures functional walking capacity by assessing the maximum distance (meters) a participant can cover in 6 minutes.
>
> **The 10 Meter Walk Test (10MWT)**[131,133] assesses functional mobility and gait by measuring the time needed to walk 10 meters at a comfortable, self-selected speed. Walking speed (m/s) is then calculated as the average of three trials.
>
> **The Stroke Self-efficacy Questionnaire (SSEQ)**[134] is a 13-item self-report scale, evaluating individuals’ confidence in performing activities of daily living. Each item is rated on a 10-point scale, where 0 indicates “not confident at all” and 10 indicates “very confident”, resulting in a total score ranging from 0 to 130.

For a subset of participants in the intervention group, we will measure **Time on Task (TOT),** defined as *“repetitive active movements performed by the participant, in line with motor learning principles, and directly related to the therapy block and personalised goals.”* Using a stopwatch, we will time the active moments for each therapy component once per week, across the three-week intervention period. Data for TOT and the process evaluation will be observed simultaneously. We will clearly distinguish between TOT and “other time.” The latter includes activities of daily living (e.g. toileting, drinking), transfer time not related to the goals, rest periods, and preparation time. Observers, who are not the treating therapists, will be positioned to avoid interfering with the therapy while maintaining sufficient visibility and audibility. A standardised method and overview for each therapy block have been developed to guide classification and timing.

##### Data analysis

We will perform an intention-to-treat analysis, including all measurements from included participants in the trial, irrespective of whether participants prematurely dropped out. Changes from baseline (T0) in FIM and SCIM scores between the experimental and control groups will be compared using a multivariate linear model with an unstructured covariance matrix, including time and treatment as main effects and a time-by-treatment interaction. The stratification variables “experience using advanced rehabilitation technology” and “pathology” will be included as fixed factors. A sensitivity analysis based on multiple imputation will be conducted if substantial missing data (>5%) are present, to assess the robustness of the results under realistic deviations from the missingness-at-random assumption. In addition, the analysis will be repeated while correcting for participant characteristics such as age, time post-stroke/SCI, and stroke/SCI severity (FIM/SCIM). The same process will be used for the secondary and stroke-specific outcome measures. Descriptive statistics will be used to characterise the usual care received by our study population, as well as to quantify the proportion of the 90-hour intervention that qualifies as Time on Task.

#### Health economic evaluation

To evaluate the cost-effectiveness of the INTeRAcT program, a trial-based cost-utility analysis will compare the costs and effects of the intervention and control groups over a nine-month time horizon post-baseline. Additionally, a Markov model will be used to estimate cost-effectiveness over a lifetime horizon.

The health economic data, including costs and effects, will be collected using a combination of methods. Baseline data will be gathered at T0. The cost data will be collected during the 9-month follow-up period through self-completion questionnaires and diaries. The diaries will be completed during the first and the ninth period of the study, while a digital questionnaire will be sent via mail monthly during the intervening months. Resource use related to the intervention will be obtained directly from the rehabilitation centre. The effects will be measured at three assessment time points: baseline (T0), after three weeks (T1), and nine months after baseline (T2), with additional data digitally surveyed each month during the nine follow-up periods (Figures 1 and 3). To reduce missing or erroneous data, the questionnaires and diaries are reviewed monthly. In case of missing or unclear information, the participant will be contacted for clarification.

##### Data: costs and utilities

This evaluation will incorporate costs and effects during the trial and follow-up period, adopting both healthcare and societal perspectives. Both perspectives will include costs covering expenses borne by health insurance as well as patient co-payments. The healthcare perspective will include all direct costs, both medical and non-medical, while the societal perspective will encompass all costs, including direct medical and non-medical costs as well as indirect costs.

**The direct medical costs** include all costs for treatment and follow-up from the healthcare perspective and all out-of-pocket contributions by the participant. The following types of resource use will be collected: primary care visits, emergency visits, inpatient stays, outpatient services, medication, diagnostic tests and medical or ADL aids.

**Direct non-medical costs** include transportation and accommodation expenses, as well as home care assistance.

**Indirect costs** comprise productivity loss (e.g. number of days away from work) and productivity losses due to informal caregiving. The productivity losses of the participants and informal caregivers will be valued using the human capital approach[135]

The effects will be expressed in **Quality-Adjusted Life Years (QALYs).** To calculate QALYs, health-related quality of life (QoL) scores are multiplied by the length of time spent in a particular health state. For example, one QALY represents one year of life in perfect health. Health-related QoL will be collected through the EQ-5D-5L questionnaire[115] and quantified in utilities derived from the national values of the EQ-5D-5L[136]. QALYs will be calculated using the area under the curve method.

To estimate the costs corresponding to the participants’ resource use, participants will be asked to provide invoices related to these expenses to ensure accurate cost reporting. Additionally, national tariffs will be used for valuation (NHIDI, BCFI, National feedback reports on hospitalisations, national cost per working hour, national cost for transportation, etc.).

##### Data analysis

First, a trial-based health economic evaluation will be conducted to assess the cost-effectiveness of the intensive, personalised rehabilitation programme compared to usual care. Individual participant data from both the intervention and control group will be used to estimate the costs and health effects of the intensive personalised rehabilitation programme compared to usual care from baseline up to nine months of follow-up. The cost-effectiveness of the intervention will be expressed in incremental cost per QALY gained, calculated as the incremental cost-effectiveness ratio (ICER):

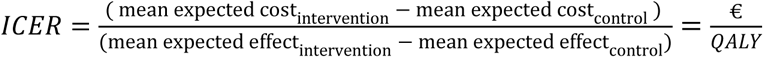

The robustness of the results will be evaluated through a deterministic and probabilistic sensitivity analysis of both costs and effects. A univariate deterministic sensitivity analysis will be conducted to assess the relative impact of individual cost components on the ICER. Results will be visually presented using tornado diagrams. A probabilistic sensitivity analysis will be performed using bootstrapping with replacement, with a minimum of 1000 iterations, to estimate the 2.5% and 97.5% percentiles of the ICER distribution. As all costs occur within a one-year timeframe, no discounting will be applied. All bootstrapped ICERs will be visualised on a cost-effectiveness plane to assess the uncertainty surrounding the ICER and the probability that the intervention is cost-effective at various willingness-to-pay thresholds. To illustrate these probabilities of acceptable ICERs, a cost-effectiveness acceptability curve will be used. In addition, exploratory subgroup analyses will be considered based on pathology, type and severity of stroke and SCI, and total rehabilitation dose.

Given the extensive data collection in this trial, some missing data may occur, therefore, appropriate data imputation techniques will be applied[137]. The analysis will be conducted using R software, following the Belgian guidelines for health economic evaluations[138], and will be reported according to the Consolidated Health Economic Evaluation Reporting Standards[139].

Secondly, we will perform a model-based health economic evaluation. Conventional practice guidelines for cost-effectiveness analysis recommend employing a time horizon that is sufficiently long to capture all relevant costs and outcomes, particularly for interventions targeting chronic conditions. In this case, a lifetime horizon as the base case analysis is often suggested[140,141]. As the absence of long-term follow-up data should not justify the failure to extend the time horizon to a relevant duration[141], a Markov model will be developed. This approach will allow us to account for the expected costs and health outcomes in both intervention and control groups beyond the follow-up period, to estimate the cost-effectiveness over a lifetime horizon. Estimates of resource use and utilities for each health state are derived from international peer-reviewed literature. Special attention will be given to the transferability of this data to the Flemish healthcare context, validated through monitoring within the research consortium. Discount rates of 3% for costs and 1.5% for utilities will be applied, following the Belgian guidelines[138]. In line with the trial-based evaluation, probabilistic sensitivity analyses will be conducted to account for uncertainty around the input parameters. R software to develop the model, calculate the incremental cost per QALY and perform the probability sensitivity analysis.

Lastly, several scenario analyses will be conducted to simulate various implementation models of the intensive programme compared to current usual care, such as providing 1–2 intensive rehabilitation boosts per year, rather than dispersing the same therapy volume across an entire year (e.g., 1-2 hours of therapy each week for a year).

A budget-impact analysis will also be performed to assess how the introduction of this intensive therapy might affect the care budgets. This analysis will consider: (1) the annual incidence of the specific population for whom this therapy is suitable; (2) the number of centres required to provide this therapy; (3) the planning timescale; and (4) the total cost of expanding this therapy across Flanders.

#### Process evaluation

The main objective of the process evaluation is to discover how complex interventions, such as INTeRAcT, function in practice and how they can be effectively transferred into clinical practice and policy when proven successful. This process evaluation will be guided by the Medical Research Council (MRC) Framework[93,142]. To gain insight into the implementation process of INTeRAcT, a mixed-methods approach will be adopted, incorporating both qualitative and quantitative methods. Data collection takes place during and until 4 weeks after the intervention (Figures 1 and 3).

The goals of the process evaluation are: (1) to explore the quality and fidelity of the implementation by examining what is delivered and how it is delivered; (2) to understand whether, how and why the therapy has an impact, by exploring both participants’ and healthcare providers’ perspectives on the program; (3) to analyse the context and the interaction between the programme and the real-world context by identifying facilitators and barriers to the implementation and sustainability of the programme, including recommendations for future implementation; (4) to assess the transferability of results by examining participants’ and therapists’ perceived experiences and impact of the intervention. The latter goal will be supported by a quantitative analysis of the data and a full health economic evaluation from a societal perspective, aiming at informing reimbursement decisions for the therapy (see previous part: health economic evaluation).

##### Data: structured diaries, semi-structured interviews and observations

**Structured therapy diaries** will provide information on how the intervention is delivered. These diaries, completed by the treating therapists, will capture data on the content of therapy, including the number and type of exercises given, the duration of therapy sessions, and other relevant treatment characteristics.

**Semi-structured interviews** will be conducted to assess the quality of the therapy as experienced by both patients and therapists. Furthermore, these interviews will explore the perceived benefit and relevance of the intervention, elicit suggestions for its implementation in routine clinical practice, assess satisfaction and acceptability by identifying facilitators and barriers, and explore stakeholders’ expectations regarding the sustainability of the programme. Therapists will also be invited to reflect on any adaptations made to the rehabilitation programme.

**Observations** will be used to gain a deeper understanding of the content and delivery of the intervention. 14 participants will be observed during therapy sessions by an unblinded researcher using a structured observation scheme. Each type of therapy block, upper and lower limb therapy (with and without technology), cardiovascular fitness training, goal-oriented training, and self-management, will be observed once per week over the 3-week intervention period. This will result in approximately 6 to 7 hours of observation per participant per week. These observations will also support the identification and interpretation of variations in outcomes across different settings and contexts.

The structured diaries, semi-structured interview guides and observation schemes were developed by the research team based on the literature[95,143–145] and expert opinions, and were piloted prior to data collection.

##### Data analysis

Descriptive statistics will be used to report the quantitative data, and analyses will be performed using IBM SPSS Statistics for Windows. Qualitative data will be analysed using both content analysis, a deductive analysis based on the defined indicators, and thematic analysis, an inductive analysis where themes will be generated from the data by the researchers using a generic qualitative approach. The aim is to interview approximately 16 participants and the 4 treating therapists to achieve meaning saturation[146], although the final sample size will be determined by saturation. Interviews will be preferably conducted face-to-face, depending on the availability of the interviewees, online interviews may also be arranged. All interviews will be audio-recorded, transcribed verbatim, and analysed using NVIVO.

To gain a more in-depth understanding of the implementation process, different types of data triangulation will be employed: (1) methodological triangulation, using a combination of interviews and observations; (2) data triangulation, incorporating perspectives from both participants and therapists; (3) theoretical triangulation, involving researchers from different disciplines to interpret the data from varied perspectives; and (4) investigator triangulation, engaging multiple researchers in the data analysis process. Throughout the research, findings will be regularly discussed within the research team.

### Ethics, data handling and dissemination

The study obtained ethical approval from the Ethics Committee Research of KU/University Hospitals Leuven, Belgium (registration number: B3222021000614, internal reference: S67164), as well as from the local ethics committee of the site, AZ Herentals, on 10 July 2023. The study will be conducted in accordance with the Declaration of Helsinki.

Throughout the clinical trial, an annual progress report (or additional reports upon request) will be submitted to the Ethics Committee, providing an overview of all (serious) adverse events that occurred during the reporting period, along with newly available safety information. The study will adhere to the approved protocol, current ICH and ICH-GCP guidelines, applicable regulatory requirements, the EU General Data Protection Regulation 2016/679 (GDPR), and relevant Belgian laws implementing the GDPR. The currently approved protocol is version 5, dated 15 March 2024. Protocol amendments will be submitted to all involved ethical committees.

Data collection, handling, processing, and transfer will be performed in compliance with applicable regulations, clinical study guidelines, and internal procedures. The details of data handling and data flow management are outlined in the study-specific Data Management Plan. Data will be pseudonymised, and (electronic) CRFs shall under no circumstances capture personal identifiers. Paper documents will be stored in a locked cupboard.

The principal investigator (PI), Geert Verheyden, is responsible for registering the Study. In addition, the PI will fulfil its ethical obligation to disseminate and make the research results publicly available. As such, the PI is accountable for the timeliness, completeness and accuracy of the reports. A variety of means will be used to disseminate the results, including academic publications, conference presentations, written reports, and communication at various stakeholder events. The full pseudonymized dataset will become available in a restricted-access repository. The dataset could be made available for other research after submitting a written request to the principal investigator, approval of an ethical committee, and based on a Data Transfer Agreement. The first article will be based on the combined outcomes of the stroke and SCI populations, covering both clinical and health economic aspects (trial- and model-based). Subsequently, publications will follow focusing on stroke-related clinical outcomes, the budget impact analysis, and the process evaluation.

## DISCUSSION

This study investigates whether the INTeRAcT motor programme is effective in improving functional independence in daily living, cost-effective, and suitable for implementation in other rehabilitation centres, compared to usual care, in individuals with chronic stroke or spinal cord injury (SCI).

This study protocol was created in close collaboration with clinical practitioners, scientific researchers, patients, and policymakers. One of the key stakeholders is the National Institute for Health and Disability Insurance (RIZIV-INAMI), Belgium’s federal agency responsible for health and disability insurance, ensuring a direct link with health policy and reimbursement discussions. Different core elements of evidence-based research were integrated in the development of this study protocol[147]. The initial inspiration for the study arose from clinical practice and was further shaped by building upon previous work by Ward and colleagues[25,95]. The protocol was refined through the incorporation of therapists’ experiences and feedback from patients.

A second strength of this project is its alignment with the phases outlined in the MRC framework[148]. This study builds on prior research and contributes to the next phase of intervention development by evaluating its clinical effectiveness, cost-effectiveness, and implementation through a structured process evaluation[142]. The broad assessment across various ICF domains ensures a holistic understanding of participants’ motor function, activities of daily living, and societal participation. In addition, the process evaluation adds value to the literature, as even promising outcomes may face challenges when translated into different rehabilitation contexts[148]. Finally, the inclusion of a health economic evaluation from both societal and healthcare perspectives will generate robust evidence on the cost-effectiveness of the INTeRAcT programme, supporting policymakers in making informed decisions regarding its broader implementation.

A third strength is the programme’s strong patient-centered approach, with therapy blocks built around individual goals and guided by a clear, structured methodology. Each therapy week offers a balanced combination of the core components: upper-limb therapy with and without advanced technology (5 hours each), lower-limb therapy with and without technology (5 hours each), goal-oriented functional training (5 hours), cardiovascular fitness training (2.5 hours), and self-management education (2.5 hours). This consistent yet adaptable structure ensures both standardisation and individualisation based on personal goals. Group-based sessions offer opportunities for peer interaction and shared learning without compromising individual needs. Furthermore, the broad geographical inclusion of participants across Belgium, supported by tailored transportation solutions, enhances the generalisability of the results and promotes equitable access.

Lastly, by evaluating the full rehabilitation programme rather than isolated components, this study adopts a whole-systems approach that more accurately reflects clinical reality. This enhances external validity and supports the integration of findings into daily practice, should the intervention prove effective.

On the other hand, certain limitations must be acknowledged. First, the intervention is delivered in a single clinical centre within the context of this study, which may limit the generalisability of the results and the replicability of the INTeRAcT programme in other settings. To mitigate this, a process evaluation is incorporated into the study to explore mechanisms of change, relevant contextual factors, and identify potential facilitators and barriers to implementation of the programme in different clinical environments[94].

Secondly, this study includes two patient populations, resulting in a heterogeneous sample. However, both populations share the common characteristic of having an acquired neurological condition in adulthood, typically presenting with similar primary symptoms such as paresis, motor and sensory impairments, and secondary complications including spasticity and cardiovascular issues. Together, these symptoms significantly affect patients’ mobility and functional independence.

Moreover, the recovery trajectories in both groups are comparable, with a critical window for motor recovery typically occurring in the first months after onset[15,16,19]. Nevertheless, evidence indicates that motor improvements remain achievable in the chronic phase, driven by exercise-dependent plasticity[20,24,25,35]. The INTeRAcT programme also incorporates principles of motor learning, which are relevant and applicable to both patient populations[15,149]. This broader inclusion enhances the generalisability of the findings to other neurological conditions, thereby increasing the relevance and applicability of the results beyond the specific populations studied.

Thirdly, as this study is designed as a pragmatic trial, the control group receives usual care, reflecting the current real-life clinical context. The inherent variability in usual care may influence clinical outcomes, therefore, both the total amount of therapy and the content delivered are documented, as outlined in the methods section.

Lastly, although the goal-directed nature of the programme provides clear source of motivation for patients[150,151], the self-enrolment procedure may attract more motivated individuals, potentially introducing selection bias. However, this phenomenon likely reflects clinical reality, where patients with higher intrinsic motivation are more inclined to participate in continued care. The process evaluation will provide crucial insights into the extent to which patient motivation contributes to the observed outcomes. Through this study, we aim to improve rehabilitation in the chronic phase of stroke and spinal cord injury. Our goals are to deliver robust evidence to support reimbursement of this motor rehabilitation package and to facilitate its implementation in other rehabilitation centres. We hope that our findings will promote an increased rehabilitation dose and intensity, the tailoring of interventions to individual patient needs, and the integration of rehabilitation technologies in the chronic phase, both in Belgium and beyond.

## Data Availability

No data have yet been generated for this study; this manuscript describes the study protocol.

## Study progress

Ethical approval was obtained by the Ethics Committee Research UZ / KU Leuven on 13 June 2023. Recruitment started in July 2023, and the first patients were randomised on 10 July 2023. The project agreement runs until September 2025, and the personal fellowship continues until October 2027. Data collection is progressing as planned.

## Author contributions

MC, MF, KP, LTT, and GV contributed to the study design, acquisition of funding, and drafting of the initial study protocol. MC, FB, MF, FJ, AS, KP, LTT, and GV were involved in the development of the trial intervention, study setup, and operational planning.

MC, MF, KP, and GV will supervise data collection and the implementation of the intervention. MC, LDB, FJ, SJ, LP, AS, and FVT are responsible for data collection, while LDB, FJ, SJ, AS, and FVT will deliver the therapy intervention.

MC, LDB, and LP drafted the original version of the manuscript. MC, MF, KP, and GV critically reviewed and edited the manuscript. All authors read and approved the final manuscript.

ChatGPT-4.5 was used to assist in rephrasing several sentences during manuscript preparation.

## Acknowledgements

We would like to thank the INTeRAcT therapists from the outpatient rehabilitation unit “To Walk Again”, AZ Herentals, Belgium, for their input in designing the INTeRAcT programme. We also extend our gratitude to the AZ Herentals team members Rudy Van Ballaer, Stefan Loots, and Daïné Lathouwers for their efforts in implementing and conducting this study protocol at the clinical site. Finally, we wish to thank Griet De Ceuster for serving as a coordinator between the KU Leuven team and AZ Herentals, and Geert Verbeke from the Leuven Biostatistics and Statistical Bioinformatics Centre for his statistical advice.

## Funding statement

The INTeRAcT trial is funded by the National Institute for Health and Disability Insurance Belgium, and a personal PhD fellowship strategic basic research of Research Foundation Flanders was awarded to Marjan Coremans (Grant no: 1SHC424N).

## Competing interest statement

The authors declare that they have no competing interests.

## Supplementary files

1. Appendix 1: Overview of demographic, socio-economic, and health-related variables
2. Appendix 2: TIDIER checklist INTeRAcT intervention

## Notes

### Competing Interest Statement

The authors have declared no competing interest.

### Clinical Trial

NCT05970367

### Author Declarations

Ethics Committee Research of KU/University Hospitals Leuven, Belgium gave ethical approval for this work. Ethics committee of the site, AZ Herentals, Belgium gave ethical approval for this work.

