## Supplementary files for "Impact of a high-dose intensive-targeted motor rehabilitation programme integrating advanced technology for adults with central neurological conditions (INTeRAcT): protocol for a single-blind randomised controlled trial with a clinical, health economic and process evaluation"

### Appendix 1 – Overview of demographic, socio-economic, and pathology-specific variables

| DEMOGRAPHIC AND SOCIO-ECONOMIC VARIABLES |
|---|
| **Self-identified sex** (M, F, X) |
| **Age** (years) |
| **Living arrangement**<br>(living with partner/and or kids, living alone, living with family, residential care facility or other) |
| **Age of cohabiting children** (years) |
| **Marital status**<br>(married, legally cohabiting, divorced, unmarried, single, widowed, or other) |
| **Education level**<br>(No formal education, primary school, lower or upper secondary education, post-secondary non-tertiary education, college graduate, university bachelor's degree, university master's degree, PhD/Doctorate, or other) |
| **Characteristics of (previous) job**<br>(job title/description; self-employed or employee) |
| **Current employment status**<br>(full-time, part-time, unemployed, retired, stay-at-home caretaker/homemaker, on disability) |
| **Self-reported monthly income** (€) |

***Appendix 2A – Demographic and socio-economic data collection.*** *Overview of the demographic and socio-economic variables and associated outcomes (indicated in parentheses) collected at baseline assessment (T0). M = male, F = Female, X = undifferentiated.*

| HEALTH-RELATED VARIABLES | |
|---|---|
| **STROKE** | **SPINAL CORD INJURY** |
| **Time since onset** (days) | |
| **Type of stroke** (ischemic, haemorrhagic, mixed) | **ASIA Impairment Scale classification** (A, B, C, D) |
| **Lesion side**<br>(left or right brain hemisphere, both or other) | **Classification of injury**<br>(complete or incomplete; paraplegia or tetraplegia / quadriplegia) |
| **Lesion location** (descriptive) | |
| **Motor hemiparesis** – **upper/lower limb** (left, right, both, not affected) | |
| **Dominant hand/leg** (left, right, unknown) | |
| **Relevant medical history** (descriptive) | |
| **Usual care** (content and frequency of physiotherapy and/or occupational therapy) | |
| **Self-reported comorbidities** (cardiovascular, respiratory, ear/nose/throat, gastrointestinal, urogenital, musculoskeletal, neurological, psychiatric, and endocrine/metabolic disorders) | |
| **Perceived impact of comorbidities on daily life**<br>(assessed on a 5-point scale ranging from “no comorbidity” to “very severe, life-threatening”) | |

***Appendix 2B – health-related data collection.*** *Overview of health-related variables and associated outcomes (indicated in parentheses) collected at baseline assessment (T0).*

**Appendix 2 – TIDieR checklist INTeRAcT intervention**

TIDieR
Template for Intervention Description and Replication

### The TIDieR (Template for Intervention Description and Replication) Checklist*:

Information to include when describing an intervention and the location of the information

| Item number | Item | Where located ** Primary paper (page or appendix number) | Other † (details) |
|---|---|---|---|
| | **BRIEF NAME** | | |
| 1. | Provide the name or a phrase that describes the intervention. | ______1_____ | _______________ |
| | **WHY** | | |
| 2. | Describe any rationale, theory, or goal of the elements essential to the intervention. | _____3-5_____ | ______________ |
| | **WHAT** | | |
| 3. | Materials: Describe any physical or informational materials used in the intervention, including those provided to participants or used in intervention delivery or in training of intervention providers. Provide information on where the materials can be accessed (e.g. online appendix, URL). | _____7-9____ | ______________ |
| 4. | Procedures: Describe each of the procedures, activities, and/or processes used in the intervention, including any enabling or support activities. | _____7-9____ | ______________ |
| | **WHO PROVIDED** | | |
| 5. | For each category of intervention provider (e.g. psychologist, nursing assistant), describe their expertise, background and any specific training given. | ______7_____ | ______________ |
| | **HOW** | | |
| 6. | Describe the modes of delivery (e.g. face-to-face or by some other mechanism, such as internet or telephone) of the intervention and whether it was provided individually or in a group. | _____7-8____ | ______________ |

| | | | |
|---|---|---|---|
| | **WHERE** | | |
| **7.** | Describe the type(s) of location(s) where the intervention occurred, including any necessary infrastructure or relevant features. | ______7_____ | _____________ |
| | **WHEN and HOW MUCH** | | |
| **8.** | Describe the number of times the intervention was delivered and over what period of time including the number of sessions, their schedule, and their duration, intensity or dose. | _____7-9____ | _____________ |
| | **TAILORING** | | |
| **9.** | If the intervention was planned to be personalised, titrated or adapted, then describe what, why, when, and how. | _____7-9_____ | _____________ |
| | **MODIFICATIONS** | | |
| **10.‡** | If the intervention was modified during the course of the study, describe the changes (what, why, when, and how). | _____n/a_____ | _____________ |
| | **HOW WELL** | | |
| **11.** | Planned: If intervention adherence or fidelity was assessed, describe how and by whom, and if any strategies were used to maintain or improve fidelity, describe them. | ____16-17____ | _____________ |
| **12.‡** | Actual: If intervention adherence or fidelity was assessed, describe the extent to which the intervention was delivered as planned. | _____n/a_____ | _____________ |

** **Authors** - use N/A if an item is not applicable for the intervention being described. **Reviewers** – use '?' if information about the element is not reported/not sufficiently reported.

† If the information is not provided in the primary paper, give details of where this information is available. This may include locations such as a published protocol or other published papers (provide citation details) or a website (provide the URL).

‡ If completing the TIDieR checklist for a protocol, these items are not relevant to the protocol and cannot be described until the study is complete.

* We strongly recommend using this checklist in conjunction with the TIDieR guide (see *BMJ* 2014;348:g1687) which contains an explanation and elaboration for each item.

* The focus of TIDieR is on reporting details of the intervention elements (and where relevant, comparison elements) of a study. Other elements and methodological features of studies are covered by other reporting statements and checklists and have not been duplicated as part of the TIDieR checklist. When a **randomised trial** is being reported, the

TIDieR checklist should be used in conjunction with the CONSORT statement (see www.consort-statement.org) as an extension of **Item 5 of the CONSORT 2010 Statement.** When a **clinical trial protocol** is being reported, the TIDieR checklist should be used in conjunction with the SPIRIT statement as an extension of **Item 11 of the SPIRIT 2013 Statement** (see www.spirit-statement.org). For alternate study designs, TIDieR can be used in conjunction with the appropriate checklist for that study design (see www.equator-network.org).
